# Validate Real-World Data-based Endpoint Measures of Cancer Treatment Outcomes

**DOI:** 10.1101/2021.06.10.21258706

**Authors:** Qian Li, Hansi Zhang, Zhaoyi Chen, Yi Guo, Thomas J George, Yong Chen, Fei Wang, Jiang Bian

## Abstract

Recently, there is a growing interest in using real-world data (RWD) to generate real-world evidence (RWE) that complements clinical trials. Nevertheless, to quantify the treatment effects, it is important to develop meaningful RWD-based endpoints. In cancer trials, two real-world endpoints are particularly of interest: real-world overall survival (rwOS) and real-world time to next treatment (rwTTNT). In this work, we identified ways to calculate these real-world endpoints with structured EHR data, and validated these endpoints against the gold-standard measurements of these endpoints derived from linked EHR and TR data. In addition, we also examined and reported the data quality issues especially the inconsistency between the EHR and TR data. Using survival model, our result showed that patients (1) without subsequent chemotherapy or (2) with subsequent chemotherapy and longer rwTTNT, would have longer rwOS, showing the validity of using rwTTNT as a real-world surrogate marker for measuring cancer endpoints.

## Introduction

In clinical trials, an endpoint is a “*precisely defined variable intended to reflect an outcome of interest that is statistically analyzed to address a particular research question*,”^1^ which is usually characterized by the type of research questions or outcomes the trials aim to assess. In cancer trials, the most common outcome-based endpoints are overall survival and measurement of tumor burden such as tumor response rate and progression free survival,^2,3^ which are usually used to measure how well a treatment worked. A number of surrogate endpoints have emerged such as time to next treatment (TTNT) that can be used as a surrogate marker for “*duration of clinical benefit*”^4,5^. Although clinical trials, especially randomized clinical trials (RCT) are considered the gold-standard to generate clinical evidence between a treatment and outcomes, they are expensive, time-consuming, and difficult to recruit sufficient participants^6,7^. Further, trial results are often not generalizable to patients who are treated in real-world settings due to issues such as overly restrictive eligibility criteria^8^. These issues are especially prevalent in cancer trials,^6,7^ where ∼97% of oncology trials ultimately fail^9^.

Recently, there is a growing interest in using real-world data (RWD) to generate real-world evidence (RWE) that complements clinical trials. The term RWD, widely promoted by the U.S. Food and Drug Administration (FDA), is referring to data collected from sources outside of conventional research settings, including electronic health records (EHRs), administrative claims, disease registries, and billing data among others^10,11^. These RWD sources contain detailed patient information such as disease status, treatment, comorbidities, and concurrent treatments that are tracked longitudinally. The information generated from RWD can provide valuable RWE about how patients are treated in real-world clinical settings, which can inform therapeutic development, outcomes research, patient care, safety surveillance, and comparative effectiveness studies^12^. Nevertheless, to quantify the treatment effects, it is important to develop meaningful RWD-based endpoints.

Another important RWD source in cancer research is the tumor registry (TR) data, which are often manually extracted from cancer patients’ medical charts (i.e. EHRs). Variables related to endpoints such as overall survival (OS), cause of death, and the date of when the first line treatment started, can be reliably obtained from TR data. However, there are a number of data gaps by using TR data alone: (1) it lacks detailed information about patients’ other characteristics (e.g. comorbidities), (2) it lacks longitudinal information about patients’ cancer treatment trajectories,^13^ and (3) it does not capture all cancer patients in the health system for various practical reasons (e.g. state and/or national TR reporting requirements and reporting delay because of the labor-intensive manual abstraction process)^14^. Nevertheless, missing patients that are not captured in TR will lead to various issues for research studies (e.g. reduced sample size leading to reduced power of the estimates). For RWD-based research studies, using linked raw EHR and TR data is ideal.

Literature on real-world endpoints is emerging and a number of endpoints have been proposed, such as real-world overall survival (rwOS), real-world time to next treatment (rwTTNT), and real-world progression-free survival (rwPFS) among others^15–17^. Nevertheless, the ability of using RWD to extract these endpoints remains to be an actively discussed area. For example, rwTTNT can be calculated based on structured EHR data alone using the dates of various procedures and diagnoses, while rwPFS will require information on the tumor itself which are often only available in unstructured clinical text (e.g. pathology reports).

Two real-world endpoints are particularly of interest: real-world overall survival (rwOS) and real-world time to next treatment (rwTTNT). The rwOS measures the duration from the date of cancer diagnosis or treatment initiation (depending on the cancer types or study aims) to the date of death or end of follow-up (or last contact); while rwTTNT measures the duration from the initiation of the first course of cancer-directed treatment to the initiation of the next line of therapy (i.e. “subsequent treatment” in the case of recurrence or progression) ^15,18^. Although rwTTNT derived from RWD sources have not been examined extensively, it can provide critical insights on real-world performance of cancer treatments^16,19^. For example, the rwTTNT can be used as a surrogate to estimate progression free survival and the effectiveness of cancer treatment^5^.

In this work, we aim to (1) identify ways to calculate real-world endpoints with structured EHR data, and (2) validate these endpoints against the gold-standard measurements of these endpoints derived from linked EHR and TR data. We focused on two real-world endpoints: rwOS and rwTTNT on early stage (stage I-III) colon cancer patients. We used RWD from the OneFlorida Clinical Research Consortium, a PCORI-funded clinical data research network contributing to the national Patient-Centered Clinical Research Network (PCORnet), with robust, longitudinal, and linked RWD for ∼15 million Floridians.

## Methods

### Overview

Our primary analysis goal of this study was two-fold: (1) to assess the alignment between EHR and TR data and describe the data gaps between the two, and (2) to assess the validity of real-world endpoints derived from EHR and TR data. To achieve these objectives, we first need to accurately depict colon cancer patients’ cancer care pathways using RWD. Conceptually, a stage I-III colon cancer patient timeline is shown in **Figure 1**.

**Figure 1.**
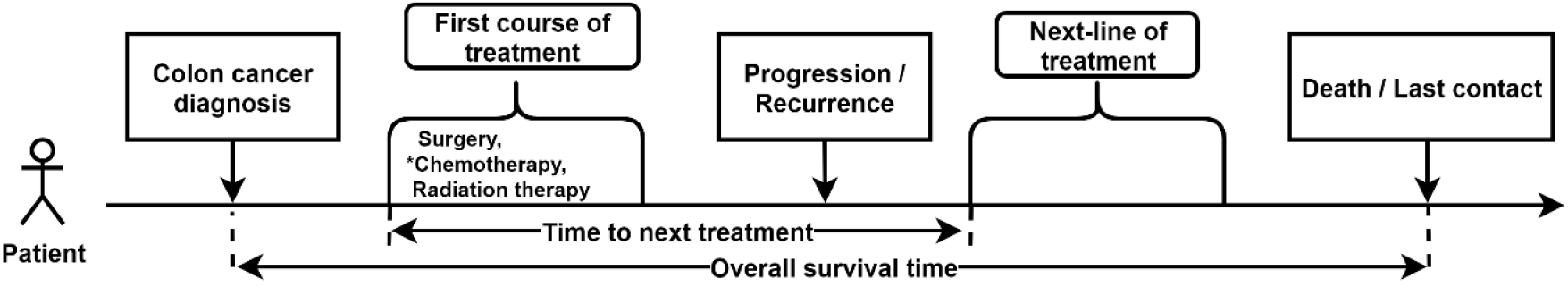
Stage I-III Colon cancer patient treatment timeline.

Our analyses involved 5 major parts, we (1) identified the events and associated dates of colon cancer diagnoses, death or last contact, and colon cancer treatments from both EHR and TR data; (2) explored the discrepancies of these events and event dates between EHR and TR data (e.g. patients identified as dead in EHR, but not in TR, and vice versa); (3) computed the rwOS; (4) identified patients’ starting points of their first course of cancer treatments and subsequent treatments, and computed the rwTTNT; and (5) examined the associations between rwOS and (i) the presence of a subsequent treatment, and (ii) rwTTNT. We hypothesize that patients who have subsequent treatment would have shorter rwOS comparing to those who do not; and for patients who have subsequent treatment, those who had a longer rwTTNT would also have a longer rwOS. Note that the first course of cancer treatment and subsequent treatment choices include surgery, radiation therapy, and chemotherapy. In this study, as we focused on stage I-III colon cancer patients, we only focused on systemic chemotherapy as depicted in clinical guidelines (see section below for details).

### Data source and study population

We used the linked EHR and TR data from the OneFlorida network. The OneFlorida network contains linked robust longitudinal patient-level RWD of ∼15 million (>60%) Floridians, including data from Medicaid claims, TR, vital statistics, and EHRs from its clinical partners. As one of the clinical research networks contributing to the national Patient-Centered Clinical Research Network (PCORnet), OneFlorida includes 12 healthcare organizations that provide care through 4,100 physicians, 914 clinical practices, and 22 hospitals, covering all 67 Florida counties. The OneFlorida data follows the PCORnet Common Data Model (CDM) including patient demographics, enrollment status, vital signs, conditions, encounters, diagnoses, procedures, prescribing and dispensing records, and lab results, etc. We extended the CDM to incorporate TR data, which follows the North American Association of Central Cancer Registries (NAACCR) standards.^20^ Currently, OneFlorida has TR data from 3 partners, which contain records of documented neoplasms (typically malignant) in their local hospital TRs. The TR records are linked with patients’ EHRs in the OneFlorida data.

The selection of the study cohort is illustrated in **Figure 2**. We first identified patients diagnosed with colon cancer in either EHR or TR data. Patients can be grouped into: (A) patients with colon cancer diagnoses in both EHR and TR data, (B) patients had colon cancer diagnoses only in the EHR data, and (C) patients had colon cancer diagnoses only in the TR data. To ensure data quality, we restricted our analyses to Group A, since it has consistent colon cancer diagnosis records from both EHR and TR data and the stage information is only available discretely in the TR data. Because stage 0 patients usually need more information to confirm their cancer diagnoses and the treatment plan for stage IV patients is more complex, we further restricted the study cohort to only patients with stage I-III colon cancer. We also excluded patients with unknown or missing stage information, and excluded patients who diagnosed with colon cancer before 2012, since our EHR data only cover patients after 2012.

**Figure 2.**
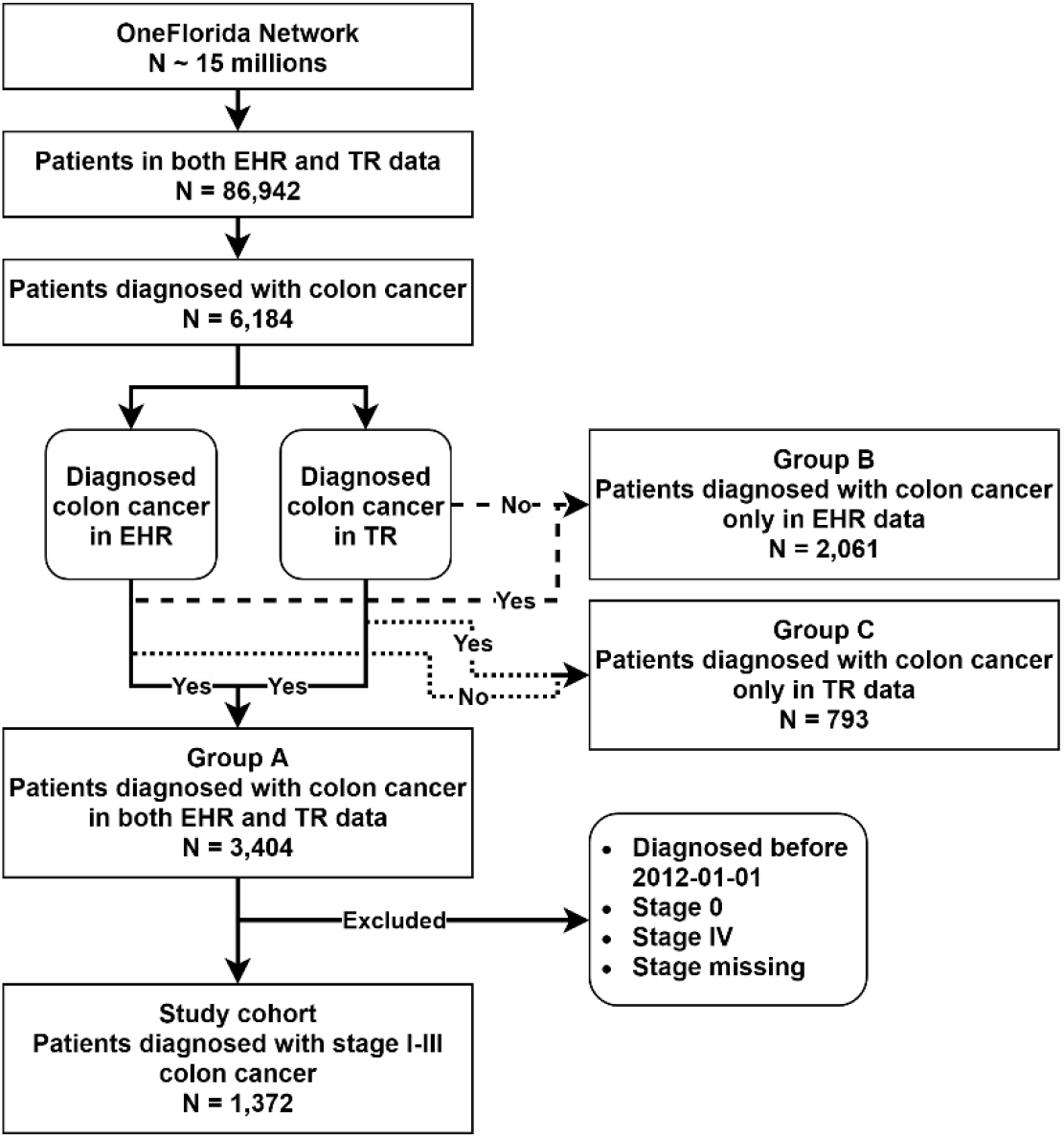
Selection process of the study cohort.

### Determination of cancer cases

To extract colon cancer patients, we used International Classification of Disease, Ninth/Tenth Revision, Clinical Modification (ICD-9/10-CM) codes 153.* and C18.* for EHR data and International Classification of Disease for Oncology-3rd edition (ICD-O-3) codes C18.0 through C18.9 for TR data. In EHR data, a colon cancer patient’s onset date was defined as the earliest encounter date with the colon cancer diagnosis.

### Determination of a patient’s last contact or death date to calculate the rwOS

We calculated rwOS as the duration from the date of the first colon cancer diagnosis to the death or last contact date. In OneFlorida, the death records in EHR data came from two sources: (1) deaths recorded by the health system (e.g. inpatient deaths or deaths reported to the health system by relatives) in patients’ EHRs, and (2) from the National Death Index (NDI), maintained by the Centers for Disease Control and Prevention (CDC) and through a privacy-preserving record linkage process using a third-party vendor, Datavant^21^. The death dates in the NDI only contain the month and year to protect privacy; thus, we imputed the death date to the first day of the month. If a patient does not have a death record, we assumed that the patient was alive. We then used the last encounter date of the patient in the EHR system as the last contact date.

On the other hand, TR data typically do contain the cancer patient’s vital status (i.e. alive or dead), and the death date (if dead), and the last contact date (if alive). However, in our TR data, the information of the last contact date is missing. Thus, to determine patients’ death or last contact date, we combined EHR and TR vital status and event dates with the following rules: (1) if patients are indicated as dead in any of the data, we treat them as dead; (2) for the dead patients, if they have death dates from both EHR and TR data, we use the one from TR data; (3) if patients are indicated as alive in both data, we treat them as alive; (4) for the alive patients, we use the last contact dates from EHR data.

### Summarization of colon cancer treatments to determine the rwTTNT

We computed rwTTNT as the duration from the beginning of the first course treatment to the beginning of subsequent treatment. To identify the colon cancer treatment, we only focused on using EHR data since TR data does not contain patients’ longitudinal treatment records. We reviewed the National Comprehensive Cancer Network (NCCN) Clinical Practice Guidelines in Oncology^22^, and identified that treatments for stage I-III colon cancer include surgery, chemotherapy, and radiation therapy. For surgery, we identified the Current Procedural Terminology (CPT) and Healthcare Common Procedure Coding System (HCPCS) codes and extracted surgery-related procedures from the EHR data. To identify chemo and radiation therapies, we started with the Cancer Therapy Look-up Tables developed by the Cancer Research Network (CRN)^23^.

As we focused on chemotherapy in this study, we first identified all chemotherapy occurrences from the EHR data and further distinguished between the first course treatment and subsequent treatments. We first defined the beginning of the first course treatment as the earliest chemotherapy record in EHR after the patient’s colon cancer diagnosis date. We implemented two rules for identifying subsequent treatments: (1) among all chemotherapy occurrences of a patient, if two adjacent chemotherapy occurrences have a gap over 90 days, we consider the former occurrence as the end of the first course treatment, and the latter occurrence as the beginning of subsequent treatment; or (2) if the patient switch to a new colon cancer treatment regimen, we consider the new regimen as the beginning of the subsequent treatment. If a patient has multiple subsequent treatments, we only count the first occurrence in our analysis.

### Statistical analyses

We first computed the confusion matrices for events (i.e., presence of colon cancer diagnosis, and vital status) obtained from EHR data vs. TR data. We computed kappa coefficients for each event from EHR and TR data. We also compared the dates of above events from EHR and TR data and reported the differences.

Further, to assess the validity of rwTTNT, we built two Cox proportional hazards models to examine the association between subsequent treatment and overall survival: (1) one, a binary variable, that considers whether the patients had subsequent treatments or not, and (2) the other that considers the rwTTNT. For both models, the outcome was the rwOS determined by both EHR and TR data. We controlled for age at diagnosis, sex, race-ethnicity, cancer stage at diagnosis, Charlson Comorbidity Index (CCI) (i.e. based on diagnosis records from EHR data^24^), and smoking status.

All data processing procedures were conducted via python and statistical analyses were performed with SAS, version 9.4 (SAS, Cary, NC, USA).

## Results

### Demographic characteristics

Our final study cohort (i.e. Group A) included 1,372 stage I-III colon cancer patients. We summarize the patients’ characteristics in **Table 1**. The mean age at colon cancer diagnosis was 65.2 years. Men and women are about the same percentage (50.7% vs 49.3%). The majority of the patients were non-Hispanic Whites (NHW). There were more stage III patients (42.6%) than stage II (33.7%) and stage I (23.7%). Most of the patients (81.0%) have no comorbidity defined by CCI. There were 41.4% non-smokers and 58.6% current smokers at baseline.

**Table 1.** Study cohort demographics characteristics.

| Demographics Characteristic (N = 1,372) | Mean / N | SD / % |
| --- | --- | --- |
| **Age at diagnosis, years** | 65.2 | 13.8 |
| **Sex** |  |  |
| Female | 676 | 49.3% |
| Male | 696 | 50.7% |
| **Race Ethnicity** |  |  |
| Non-Hispanic White | 821 | 59.8% |
| Non-Hispanic Black | 230 | 16.8% |
| Hispanic | 198 | 14.4% |
| Other | 123 | 9.0% |
| **Colon Cancer Stage at diagnosis** |  |  |
| I | 325 | 23.7% |
| II | 462 | 33.7% |
| III | 585 | 42.6% |
| **Charlson Comorbidity Index at diagnosis** |  |  |
| None (0) | 1112 | 81.0% |
| Mild (1-2) | 117 | 8.5% |
| Moderate (3-4) | 62 | 4.5% |
| Severe ($\geq 5$) | 81 | 5.9% |
| **Smoking status at diagnosis** |  |  |
| Never | 568 | 41.4% |
| Ever | 804 | 58.6% |

### Cancer diagnosis comparison

All 1,372 patients had diagnosis records of colon cancer from both EHR and TR data; however, there are differences in the diagnosis dates between EHR and TR data as shown in **Table 2**. 73.4% of the differences were in less than 1 month. There were more patients who had TR diagnosis dates earlier than EHR. 16.2% of the difference were in their TR records with 1 to 3 months earlier than EHR and 9.0% more than 3 months earlier. There were very few differences in which diagnoses in EHR were earlier than TR, 0.9% were 1 to 3 months earlier and 0.5% were more than 3 months earlier.

**Table 2.** Colon cancer diagnosis date difference between EHR and TR data.

| Colon cancer diagnosis date difference (N = 1,372) | Frequency | Percent |
| --- | --- | --- |
| EHR record is 1-3 month earlier than TR record | 12 | 0.87% |
| EHR record is > month earlier than TR record | 7 | 0.51% |
| TR record is 1-3 month earlier than EHR record | 222 | 16.18% |
| TR record is > month earlier than EHR record | 124 | 9.04% |
| Within 1 month | 1007 | 73.40% |

Even through our statistical analyses focused on Group A patients, we further explored patients’ cancer diagnosis records in Group B and Group C as shown in **Table 3**. For patients in Group B (i.e. patients had colon cancer diagnoses only in the EHR data), more than 50% (N = 1,051) of the patients were diagnosed with rectum cancer. For patients in Group C (i.e., patients had colon cancer diagnoses only in the TR data), 75% (N = 602) of the patients’ first cancer diagnoses were before 2012, which would not be captured in our EHR data. For the rest of 25% (N = 195) patients, many of their first cancer diagnoses was secondary malignancy.

**Table 3.**
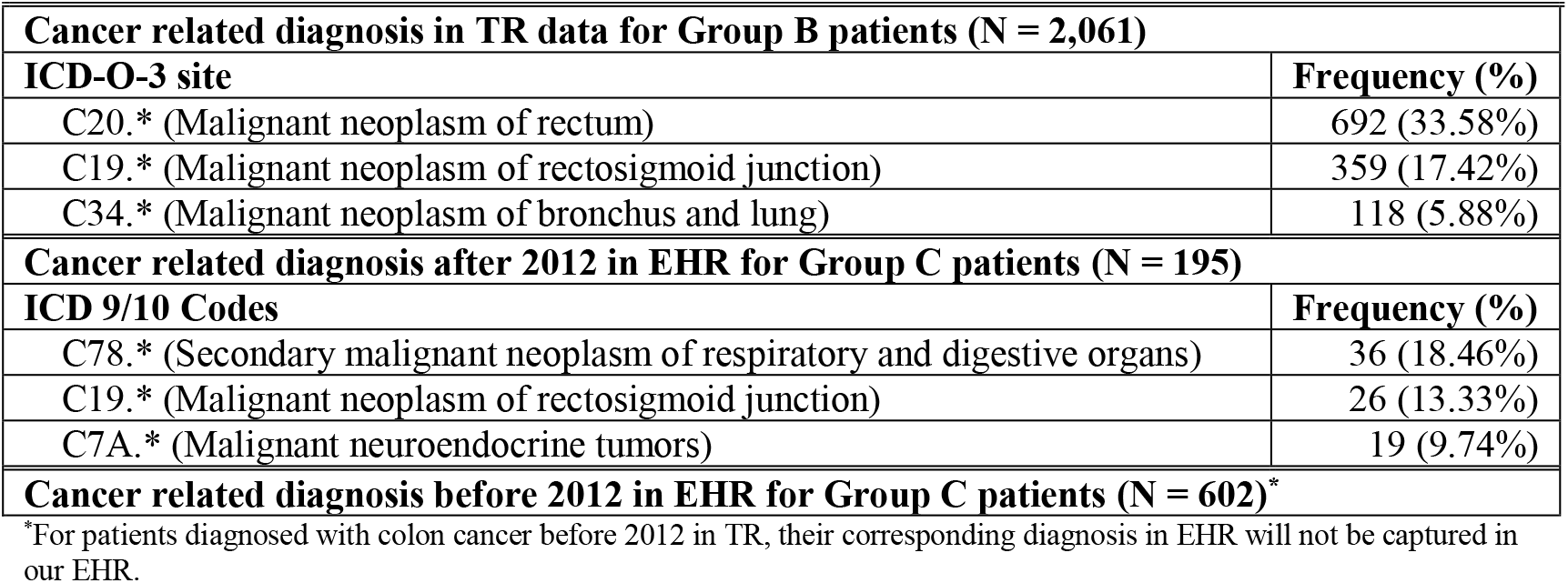
Cancer related diagnosis for Group B and C patients.

### Vital status and death date comparison

For Group A, in EHR data, there were 248 (18.08%) patients with death information; while 272 (19.83%) patients were deceased in the TR data. The confusion matrix is shown in **Table 4**. The Kappa coefficient was 0.66 (95% CI: 0.61 - 0.71). 75.8% of patients were identified as alive and 13.7% of patients were identified as dead in both data. There was a small portion of patients with inconsistent vital status across these data, 4.37% alive in TR but deceased in EHR and 6.12% deceased in TR but alive in EHR.

**Table 4.** Confusion matrix for vital status in EHR and TR data.

| Frequency<br>Percent | TR vital status |  |  |
| --- | --- | --- | --- |
|  | Alive | Dead | Total |
| EHR vital status |  |  |  |
| Alive | 1040<br>75.8% | 84<br>6.12% | 1124<br>81.92% |
| Dead | 60<br>4.37% | 188<br>13.7% | 248<br>18.08% |
| Total | 1100<br>80.17% | 272<br>19.83% | 1372<br>100% |

We compared the death dates for those recorded as deceased in both EHR and TR data. Among those 188 patients, only one patient has the death dates that are different between TR and EHR data over 1 month; difference for others are all within 1 month.

**Figure 3** shows the distribution of rwOS in months. There are 5 patients with negative rwOS values, where their death or last contact dates were earlier than their colon cancer diagnosis dates. This is likely due to data entry errors or other data quality related issues. We removed these patients with negative rwOS for the statistical modeling. The maximum length of rwOS is 101 months and the median is 22.6 months.

**Figure 3.**
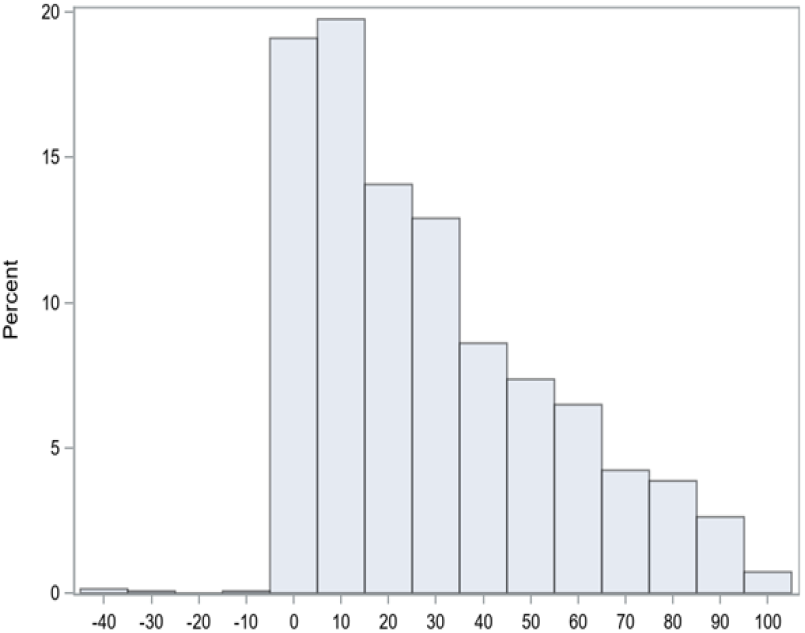
Distribution of real-world overall survival in months.

With EHR data, we identified 174 (12.68%) patients who had subsequent treatments. **Table 5** shows the presence of subsequent treatment across different cancer stages. Among stage I patients, 5.85% of them had subsequent treatments. While in stage II and stage III patients, the percentages raise up to 10.82% and 17.95%, respectively. The distribution of rwTTNT is shown in **Figure 4**. The average time to next treatment is 9.1 months with a standard deviation of 10.6 months and the median is 6.3 months.

**Table 5.** Presence of subsequent treatment by colon cancer stage.

| Colon Cancer Stage at diagnosis | Presence of Subsequent Treatment |  |  |
| --- | --- | --- | --- |
|  | No | Yes | Total |
| **I** | 306<br>94.15% | 19<br>5.85% | 325 |
| **II** | 412<br>89.18% | 50<br>10.82% | 462 |
| **III** | 480<br>82.05% | 105<br>17.95% | 585 |
| **Total** | 1198 | 174 | 1372 |

**Figure 4.**
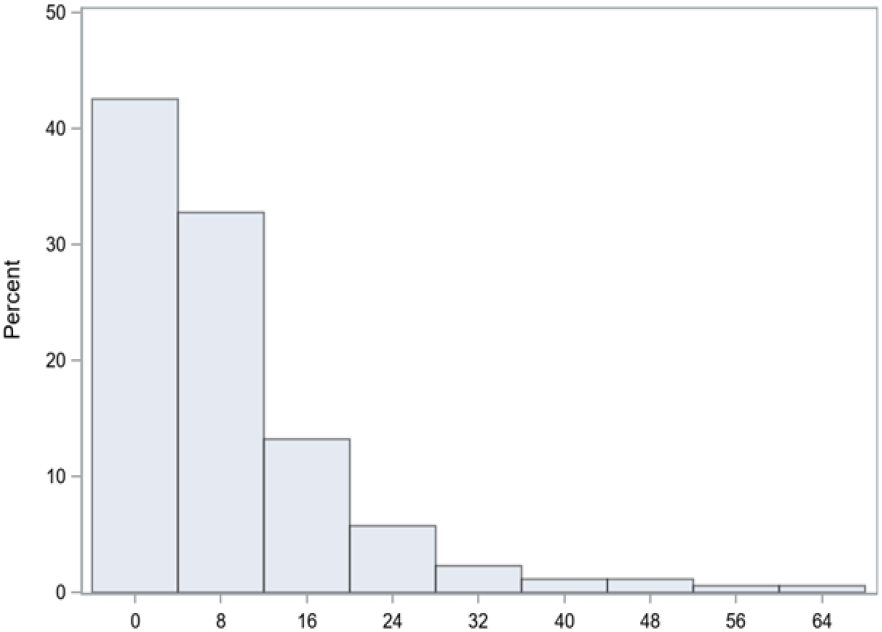
Distribution of real-world time to next treatment in months.

### Association between presence of subsequent treatment / rwTTNT and rwOS

**Table 6** shows the estimates of Cox proportional hazards model on rwOS using the presence of subsequent treatments as a predictor. Age at diagnosis is significantly associated with rwOS, with a hazard ratio (HR) of 1.038. Increasing in age at diagnosis would shorten rwOS. Sex is not a significant predictor for rwOS; but the HR is 1.246, meaning males have shorter rwOS compared to females. Compared to non-Hispanic Whites, non-Hispanic Blacks and Hispanic have lower hazards, with HRs of 0.659 and 0.623, respectively. Cancer stage at diagnosis is significantly associated with rwOS, stage I and II compared to stage III have much longer rwOS, with HRs of 0.454 and 0.517, respectively. Patients with CCI > 0 have shorter rwOS compared to those that are not. The smoking status at baseline is not significantly associated with rwOS. Patients with subsequent treatment have a higher hazard than those without (HR: 1.203). Present of subsequent treatment leads to a shorter rwOS, but it is not statistically significant (p = 0.2173).

**Table 6.** Estimates of real-world overall survival with presence of subsequent treatments as a predictor.

| Parameter |  | Hazard Ratio | 95% Hazard Ratio Confidence Limits |  | p Value |
| --- | --- | --- | --- | --- | --- |
| Age |  | 1.038 | 1.028 | 1.048 | <.0001+ |
| Sex | Male vs Female | 1.246 | 0.997 | 1.556 | 0.0530# |
| Race-ethnicity | NHB vs NHW | 0.659 | 0.473 | 0.918 | 0.0138+ |
| Race-ethnicity | Hispanic vs NHW | 0.623 | 0.425 | 0.915 | 0.0157+ |
| Race-ethnicity | Other vs NHW | 0.911 | 0.600 | 1.383 | 0.6614 |
| Colon cancer stage at diagnosis | I vs III | 0.454 | 0.332 | 0.619 | <.0001+ |
| Colon cancer stage at diagnosis | II vs III | 0.517 | 0.399 | 0.670 | <.0001+ |
| Charlson comorbidity index at diagnosis | > 0 vs = 0 | 1.724 | 1.343 | 2.214 | <.0001+ |
| Smoking status at diagnosis | Ever vs Never | 1.027 | 0.805 | 1.311 | 0.8288 |
| Presence of subsequent treatment | Yes vs No | 1.203 | 0.897 | 1.614 | 0.2173 |

+: statistically significant at 0.05 level; #: close to statistically significant at 0.05 level.

**Figure 5** shows the Kaplan-Meier curves on rwOS, strata by sex, race-ethnicity, stage, CCI, and presence of next treatment. Males have lower survival probabilities than females. Patients with subsequent treatment have lower survival probabilities at later phase.

**Figure 5.**
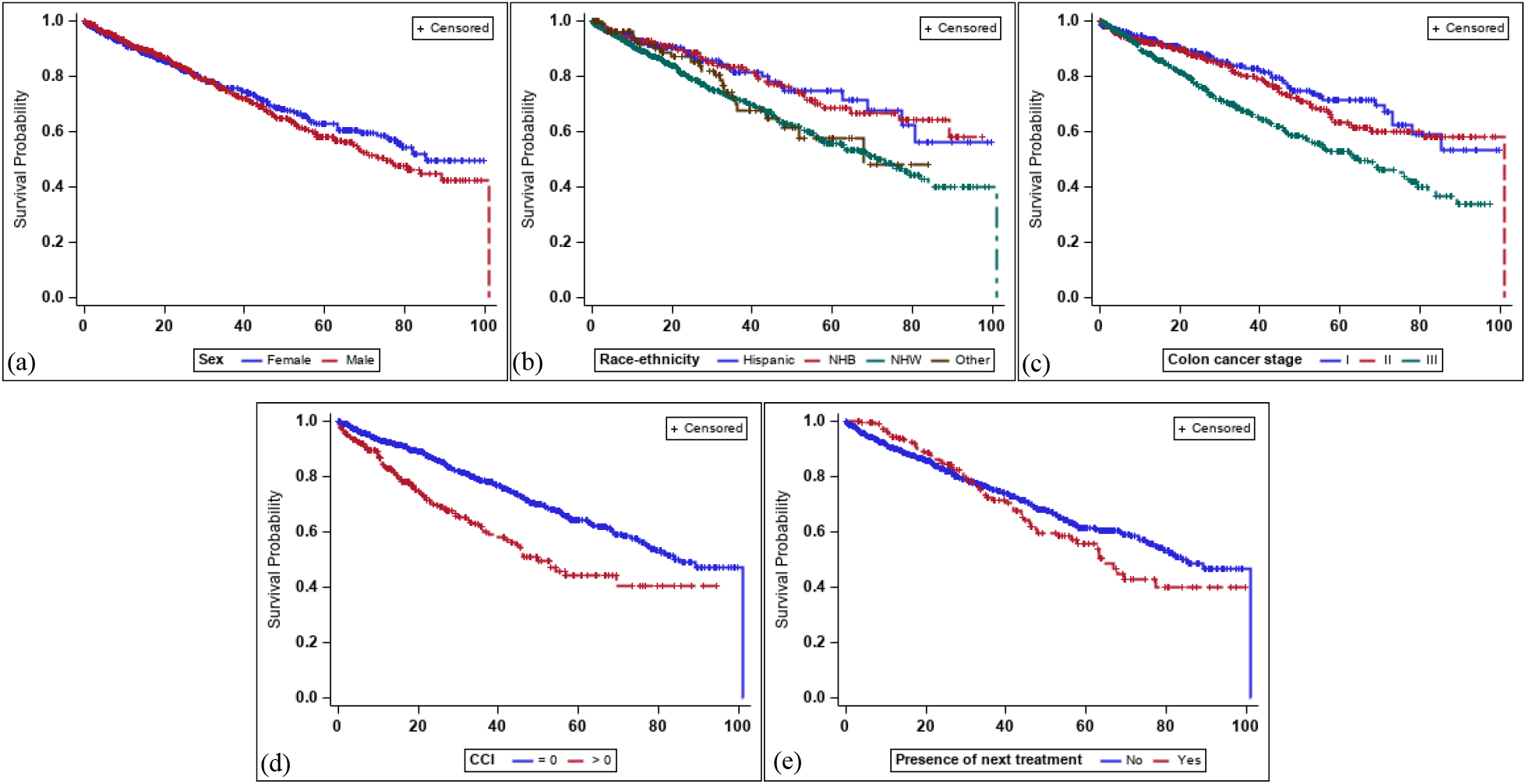
Kaplan-Meier survival plots on real-world overall survival strata by (a) sex, (b) race-ethnicity, (c) colon cancer stage, (d) Charlson comorbidity index (CCI), and (e) presence of next treatment.

**Table 7** shows the estimates of Cox proportional hazards model on rwOS with rwTTNT as a predictor on patients who had subsequent treatments. rwTTNT itself is statistically significant, having a HR of 0.973, which means patients with longer rwTTNT would have a longer rwOS.

**Table 7.**
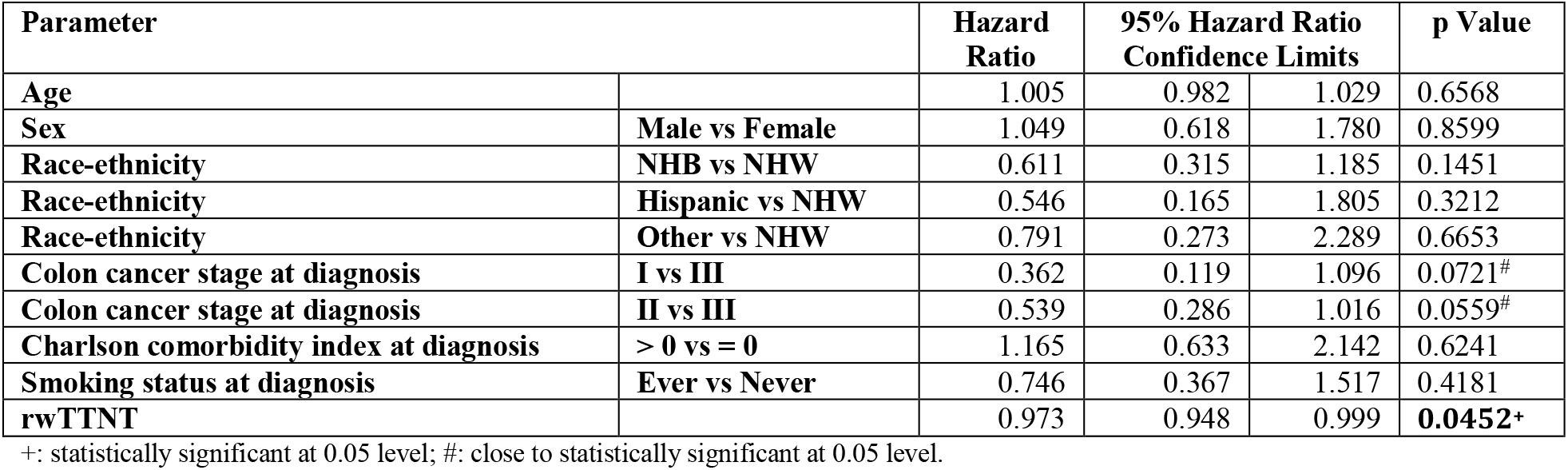
Estimates of Cox proportional hazards model for real-world overall survival with real-world time to next treatment as a predictor.

| Parameter |  | Hazard Ratio | 95% Hazard Ratio Confidence Limits |  | p Value |
| --- | --- | --- | --- | --- | --- |
| Age |  | 1.005 | 0.982 | 1.029 | 0.6568 |
| Sex | Male vs Female | 1.049 | 0.618 | 1.780 | 0.8599 |
| Race-ethnicity | NHB vs NHW | 0.611 | 0.315 | 1.185 | 0.1451 |
| Race-ethnicity | Hispanic vs NHW | 0.546 | 0.165 | 1.805 | 0.3212 |
| Race-ethnicity | Other vs NHW | 0.791 | 0.273 | 2.289 | 0.6653 |
| Colon cancer stage at diagnosis | I vs III | 0.362 | 0.119 | 1.096 | 0.0721<sup>#</sup> |
| Colon cancer stage at diagnosis | II vs III | 0.539 | 0.286 | 1.016 | 0.0559<sup>#</sup> |
| Charlson comorbidity index at diagnosis | > 0 vs = 0 | 1.165 | 0.633 | 2.142 | 0.6241 |
| Smoking status at diagnosis | Ever vs Never | 0.746 | 0.367 | 1.517 | 0.4181 |
| rwTTNT |  | 0.973 | 0.948 | 0.999 | **0.0452<sup>+</sup>** |

+: statistically significant at 0.05 level; #: close to statistically significant at 0.05 level.

**Figure 6** shows the Kaplan-Meier survival plots on rwOS for the patients with subsequent treatment only, strata by sex, race-ethnicity, stage, and CCI. Because of small sample size, the difference between the survival curves for each stratum, except the colon cancer stage, are small.

**Figure 6.**
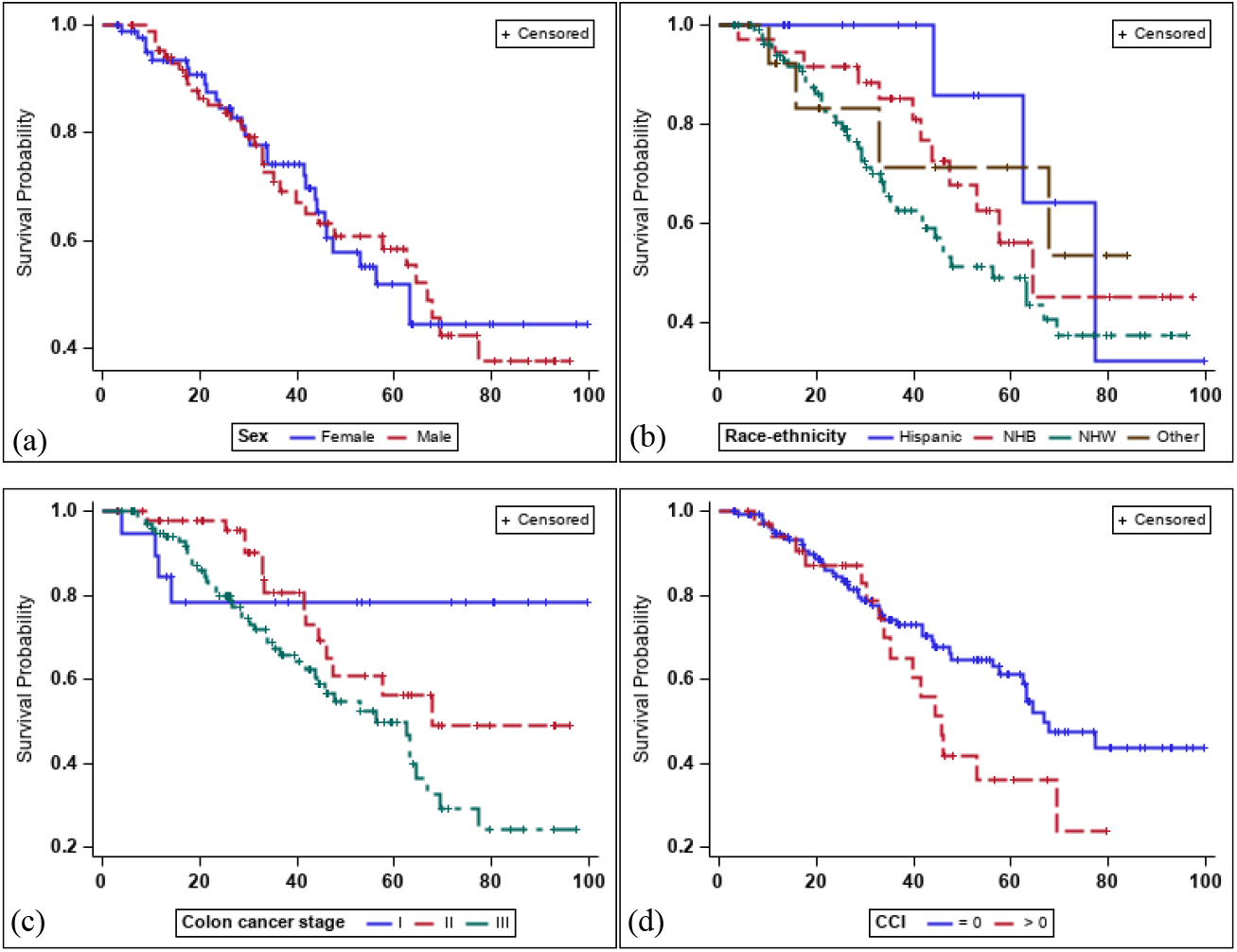
Kaplan-Meier survival plots on real-world overall survival for the patients with subsequent treatment only, strata by (a) sex, (b) race-ethnicity, (c) colon cancer stage, and (d) Charlson comorbidity index (CCI).

## Discussion and Conclusion

In this study, we identified and validated two real-world endpoints: rwOS and rwTTNT in stage I-III colon cancer patients. Using RWD from linked EHR and TR, we identified the necessary events on colon cancer patients’ treatment timelines, including (1) cancer diagnosis, (2) vital status and last contact date, and (3) cancer treatments, to established the two real-world endpoints. We assess the discrepancy for these events and related dates between EHR and TR data. With longitudinal records in EHR data, we differentiate the first course of cancer treatment (focusing on chemotherapy) and the subsequent next-line treatment, which were used to compute rwTTNT. We showed that the patients with subsequent treatment would have shorter overall survival, although it is not statistically significant. We also showed that increase in rwTTNT would increase rwOS (i.e. a longer time to next treatment leads to a longer overall survival time).

There are discrepancies between our EHR and TR data, revealing various potential data quality issues with RWD. First of all, the diagnosis records of colon cancer do not line up between the EHR and TR data; there are patients who had colon cancer diagnosis in only EHR (Group B) and in only TR (Group C). Possible reasons for such discrepancy between EHR and TR could be (1) patients with rectal cancer was misdiagnosed as colon cancer in the first place, but later confirmed as rectal cancer and reported to TR; (2) TR has a significant reporting latency (i.e. typically more than 6 months delay in reporting) because of the manual abstraction process, and (3) patient continuity issues (i.e. patients seeking care across different health systems) that leads to EHR data continuity issues. The difference in the dates of cancer diagnosis between EHR and TR is better than we expected. For those patients with consistent colon cancer diagnoses in both datasets (i.e. Group A, our final study cohort), over 70% of the patients have a difference in the diagnosis dates less than 1 month between the EHR and TR data. Further, we did not expect that the last contact date information for alive patients is missing in our TR data. Nevertheless, the difference between EHR death date and TR death date is negligible, where only 1 patient has different deceased dates over 1 month. Although RWD have data quality issues in completeness and accuracy, linking RWD from multiple sources such as EHR and TR could provide a more accurate depiction of the patients’ care and health status.

In terms of real-world endpoints, past studies like Stewart et.al 2019^15^ have only shown a positive correlation (simply Spearman’s rank-order correlation) between rwTTNT and rwOS without controlling for other covariates. In our study, we built two Cox proportional hazard models for survival analysis and controlled for a number of important risk factors of rwOS such as age, gender, race-ethnicity, cancer stage, and CCI. In the first model, we dichotomized the rwTTNT as having vs. not having next-line treatment, which led to a larger sample size. Thus, we have more statistical power to model and extent the dimension of the relation between rwTTNT and rwOS. In the second Cox model, we modeled the relation between the actual length of the rwTTNT and patients’ overall survival and yielded a statistically significant result. As Cox model is commonly used in modeling clinical trial results; being able to run Cox models with real-world endpoints and RWD is significant, such that RWD-based analyses can generate results compatible to clinical trials. Being able to generate validated surrogate markers such as the rwTTNT that we investigated in this study, would provide health outcomes and comparative effectiveness research investigators a new tool to leverage the large collections of RWD that is being increasingly available.

Our study is not without limitations. First, our sample size for modeling rwTTNT and rwOS is relatively small. As additional sites contribute their TR data to OneFlorida, we can expand the study cohort to achieve higher statistical power. Second, cancer stage information is only available discretely in our TR data currently, however, are prevalent in unstructured documents stored in EHRs (e.g. pathology reports). We shall explore advanced natural language processing (NLP) tools to unlock the critical information such as cancer stage and other tumor characteristics, especially if only EHR data are available. Third, current rules for identifying the first course of treatment and subsequent treatments can be improved. For example, our rule defines a 90-day wash-out period to differentiate the end of the first course treatment and beginning of the subsequent treatment. We assumed that the 90-day gap could eliminate the effects of the previous treatment. However, different drugs have different wash-out period, leading to potential misclassification of the different treatment courses. We can potentially further decompose the rules for each type of colon cancer chemotherapy. For example, some trials^25^ defined a 1-month wash-out period to eliminate the effects of the previous treatment for colon cancer. More fine-grained rules and/or computable phenotypes should be developed and validated for the future studies.

In summary, we identified rwTTNT and rwOS from EHR and TR data. We assessed the alignments between EHR and TR data on the colon cancer diagnosis, treatment, and survival related events. We showed that rwTTNT is positively correlate with rwOS. Thus, rwTTNT could be an important surrogate marker for studies that utilized RWD and ultimately help to optimize colon cancer treatment to extent overall survival.

## Data Availability

OneFlorida data can be requested at https://onefloridaconsortium.org/front-door/; Since OneFlorida data is a HIPAA limited data set, a data use agreement needs to be established with the OneFlorida network.

## Acknowledgment

This work was supported in part by NIH grants R01CA246418, R21AG068717, and R21CA245858 and the OneFlorida Clinical Research Consortium (CDRN-1501-26692) funded by PCORI. The content is solely the responsibility of the authors and does not necessarily represent the official views of the NIH or PCORI.

